# A Multi-Component Telehealth Intervention to Improve Oncofertility Care Delivery among Young Cancer Patients: A Pilot Study

**DOI:** 10.1101/2022.02.18.22271158

**Authors:** Emily Yang, Anna Dornisch, Laura Nerb, Teresa Helsten, Bonnie N. Kaiser, Paula Aristizabal, Saro Armenian, Lilibeth L. Torno, Nicole M. Baca, Mark C. Genensen, H. Irene Su, Sally A. D. Romero

## Abstract

**Purpose:** Oncofertility care for pediatric, adolescent, and young adult cancer patients remains under-implemented across adult and pediatric oncology settings. We pilot tested an electronic health record (EHR)-enabled multi-component oncofertility intervention (including screening, referral, and fertility consult) in an adult academic oncology program and systematically assessed intervention fit to pediatric and community oncology programs.

**Methods:** Using surveys (n=33), audits (n=143), and interviews (n=21) guided by implementation science frameworks, we pilot tested the EHR-enabled intervention for oncofertility care in young cancer patients at an adult oncology program and evaluated implementation outcomes. We interviewed healthcare providers from seven regional oncology and fertility programs about intervention fit to their clinical contexts.

**Results:** We recruited 33 healthcare providers from an adult oncology setting and 15 healthcare providers from seven additional oncology and fertility settings. At the adult oncology setting, the intervention was found to be appropriate, acceptable, and feasible and improved the screening of fertility needs (from 30% pre- to 51% post-intervention), yet some patients did not receive appropriate referrals to fertility consults. Providers across all settings suggested content and context modifications, such as adding options to the intervention or allowing the screening component to pop up at a second visit, to improve and adapt the intervention to better fit their clinical care contexts.

**Conclusions:** We found that the EHR-enabled intervention increased the rate of goal-concordant oncofertility care delivery at an adult oncology program. We also identified facilitators, barriers, and needed adaptations to the intervention required for implementation and scaling up across diverse oncology settings.

## Introduction

Pediatric, adolescent, and young adult cancer patients undergo radiation, chemotherapy, surgery, and/or endocrine therapy that may harm future fertility.^1,2^ Clinical guidelines from oncology and fertility societies recommend oncofertility care, specifically that healthcare providers discuss infertility risk with all reproductive-aged patients and offer appropriate fertility preservation options or referrals to reproductive specialists for interested patients.^3-6^ Despite these longstanding clinical guidelines, oncofertility care is not routinely provided in pediatric and adult oncology settings due to multi-level barriers, such as incomplete patient-provider communication,^7^ lack of clear referral pathways,^8-10^ limited access to fertility programs,^11^ and parental unwillingness to discuss reproduction with children.^12^

A consistent theme across these multi-level barriers is the need to systemize processes for clinics and providers to improve routine patient engagement in oncofertility care. Interventions to address these barriers within health systems are limited in initial design to systematically address multi-level barriers, evaluation of efficacy, and scaling beyond single institutions. Most oncofertility care programs are single-institution and multi-component to include patient education conducted by oncology or fertility healthcare providers and referral from oncology to fertility.^7,13-21^ Program components vary but include provision of oncology provider education, resource-intensive oncofertility navigators, electronic health record (EHR)-enabled screening of eligible patients, decision support, and psychological support. A few programs developed their interventions systematically using implementation or improvement science methodology,^18,19^ while most did not specify and are thus at risk of barrier-intervention mismatch. Further, no scaling up of oncofertility care beyond single institutions has been undertaken.

Guided by the Consolidated Framework for Implementation Research (CFIR),^22^ an implementation science framework, our group systematically identified three key steps for implementing oncofertility care: 1) needs screen for all young cancer patients, 2) referral to fertility specialists (as needed), and 3) fertility specialists consultation and FP services (as needed).^21^ Additionally, we found that EHR systems use functionalities that standardize, tailor, and minimize steps in clinical workflows; these functionalities are shareable between health systems and scalable. We also identified telehealth to support gaps in delivering and accessing oncofertility care. Through provider and patient stakeholder engagement, we leveraged both EHR-enabled systems and telehealth functionalities to develop a scalable multi-component intervention that encompasses an oncofertility needs screening, referral from oncology provider to fertility specialist, and oncofertility counseling. The objectives of this study were two-fold: 1) pilot test the multi-component intervention for oncofertility care at three outpatient adult oncology clinics and 2) evaluate intervention fit at additional oncology programs in the region.

## Methods

### Participants

Between March-October 2020, we recruited, consented, and enrolled healthcare providers (physicians and advanced practice providers [APPs]) from three outpatient adult oncology clinics at the University of California San Diego (UCSD) Moores Cancer Center to participate in surveys and semi-structured interviews about the pilot multi-component oncofertility intervention. Participants included members of the breast, urology, and hematology oncology teams.

Between June-September 2020, we recruited, consented, and enrolled oncology providers, social workers, and fertility providers from regional academic and community adult and pediatric oncology and fertility programs to participate in semi-structured interviews focused on evaluating the intervention for fit to their clinical contexts. Participating sites included Children’s Hospital of Orange County, City of Hope, Cedars Sinai, Rady Children’s Hospital, University of California Los Angeles, Eisenhower, and Kaiser Southern California. Participants included individuals nominated for having expertise in pediatric and adolescent solid and hematologic malignancies, survivorship, and female and male fertility preservation.

The study was approved by the Institutional Review Board at UCSD.

### EHR-enabled Multi-Component Intervention

The intervention included three core components facilitated by the EPIC EHR system: 1) oncofertility needs screen using a best practice advisory (BPA), 2) BPA-linked oncofertility referral pathway from oncology to fertility, and 3) oncofertility counseling with fertility specialists (Figures 1 and 2). The BPA and BPA-linked referral order were tailored for the clinical setting at the adult oncology program (Table 1). The BPA was designed to prompt oncology providers (physicians and APPs) that oncofertility counseling for newly diagnosed and post-treatment cancer survivors is recommended; fires as a pop up when an oncology diagnosis is entered at new patient visits for female patients ages 0 to 42 years and male patients ages 0 to 50 years; and has an embedded fertility referral defaulted to order. The provider can accept the referral order, choose not to place an order and select a reason for not referring, or dismiss the BPA. The referral order was designed to be linked to the BPA to reduce oncology provider search effort, to require key cancer treatment and timeframe data to facilitate communication between oncology and fertility, and to be placed in a ‘STAT’ dedicated work queue overseen by fertility clinic staff for insurance authorization and scheduling. The provider’s response to the BPA was linked to backend logic to reduce reminder fatigue. Referrals not placed for reasons of no fertility needs, poor prognosis, or not enough time before treatment turns off the BPA for all eligible providers for two years. Referral not placed due to patient’s declining to address the issue in that visit would trigger a ‘snooze’ and enable the BPA to pop up at the follow up visit with that provider or in a new visit with another oncology provider. Oncofertility consultations were offered in-person or via telehealth, and telehealth video visits were conducted through the EPIC patient portal MyChart.

**Table 1.**
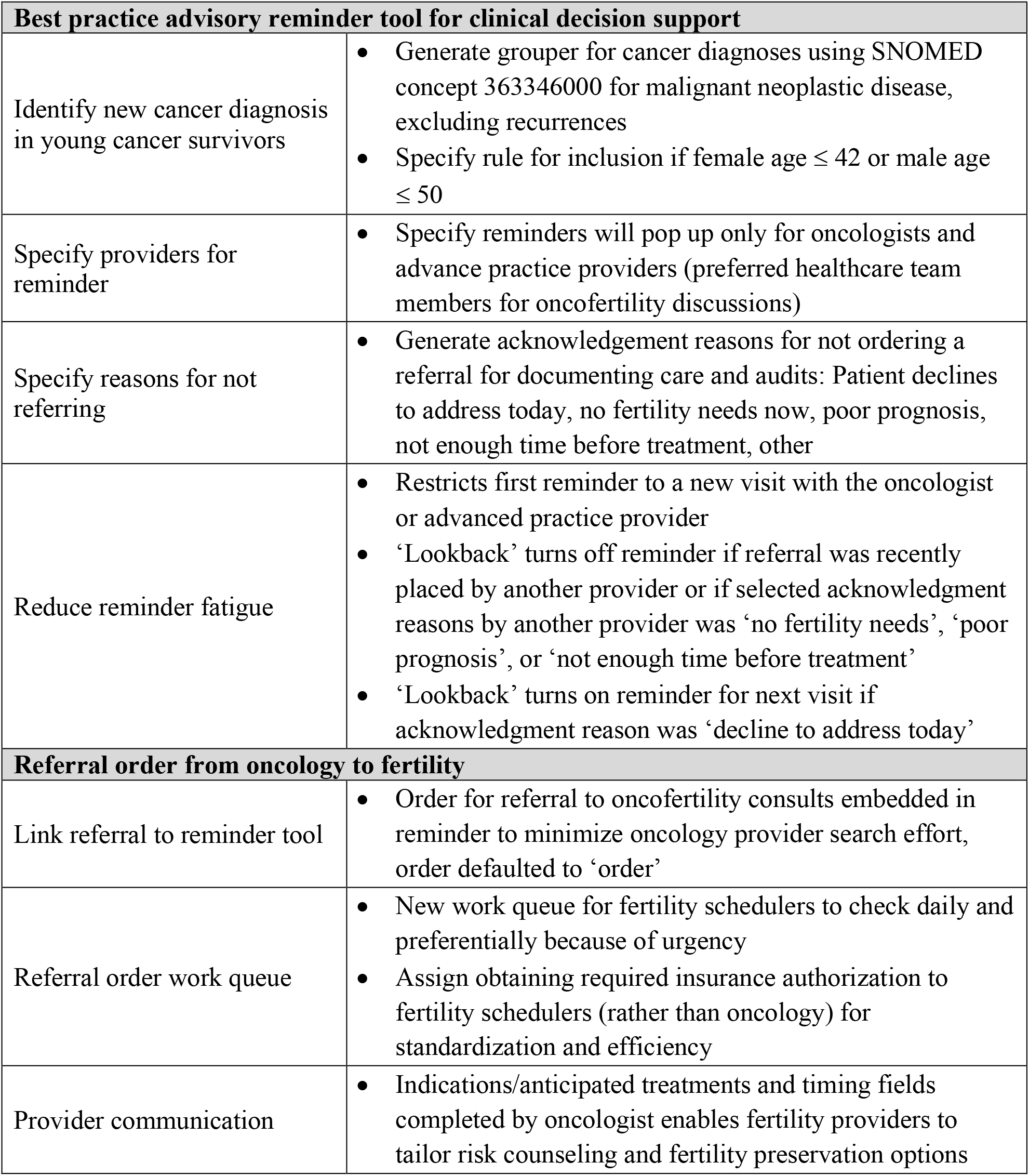
Oncofertility needs screen and referral pathway in EPIC EHR system: Example design decisions and specifications

**Figure 1.**
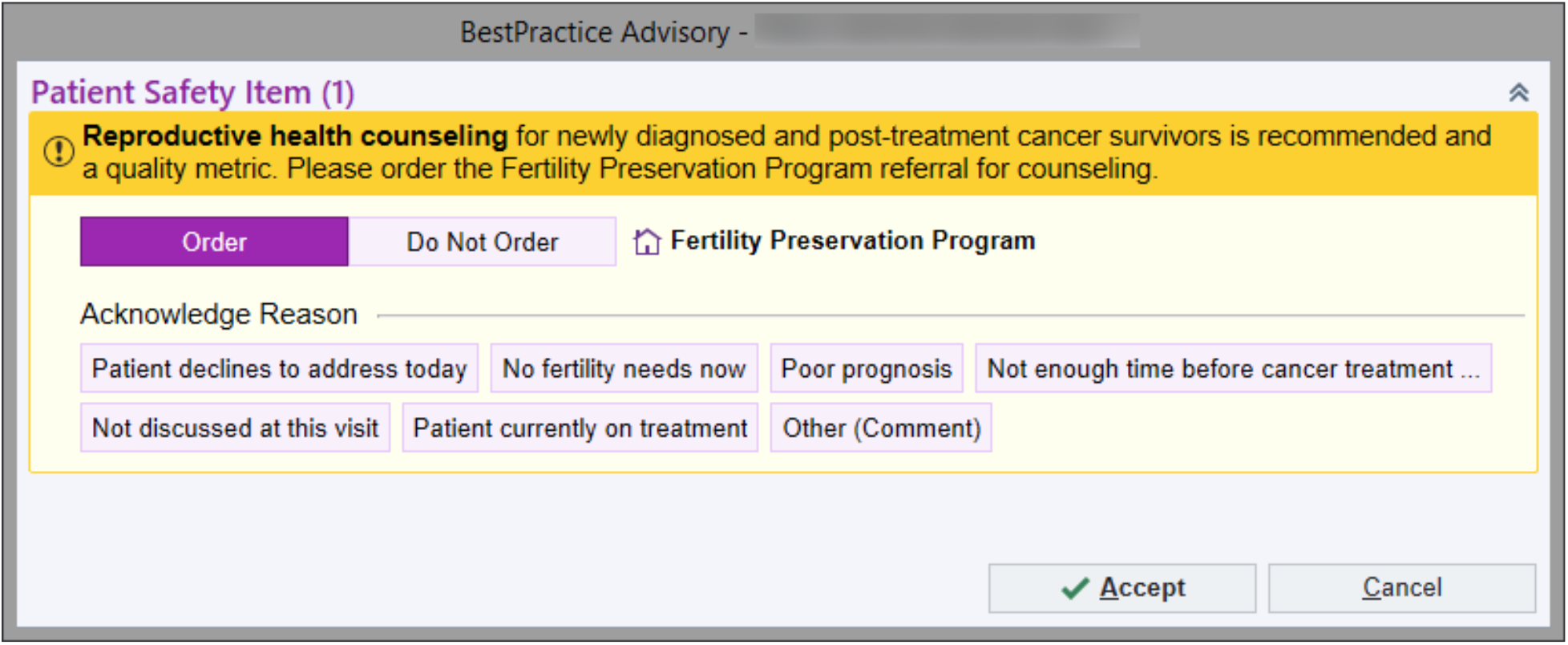
Electronic health record-enabled Best Practice Advisory oncofertility needs screen and referral pathway to oncofertility consultation.

**Figure 2.**
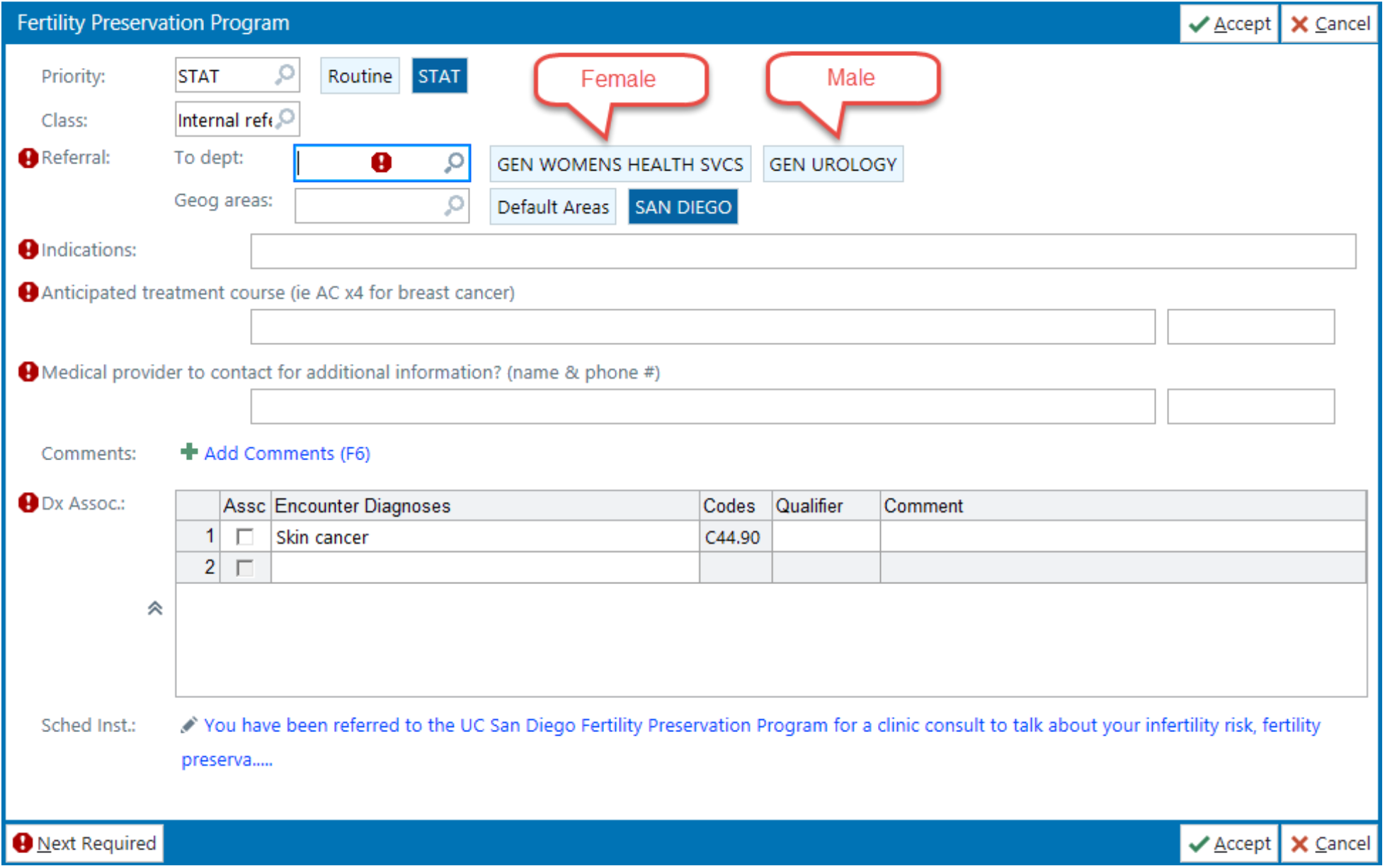
Electronic health record-enabled Best Practice Advisory linked referral order for oncofertility consultation.

Implementation was supported by two strategies: 1) a 15-minute provider educational session to discuss guideline-recommended oncofertility care at cancer diagnosis, introduce the intervention, and describe associated workflow prior to implementation; and 2) EHR audit report based on the BPA and referral order outcomes. The intervention was activated in April 2020 and pilot tested for three oncology teams (breast, urology, and hematology) at UCSD Moores Cancer Center.

### Surveys

Prior to and three months post-implementation at UCSD, we recruited a purposive sample of providers to complete questionnaires on demographic characteristics and feasibility, acceptability, and appropriateness of intervention components.^23^ Each measure consisted of four questions asking participants how much they agreed with each statement, using a five-point Likert scale (Strongly Disagree [1] to Strongly Agree [5]).

### Interviews

We conducted semi-structured video call interviews with providers from UCSD and additional sites. Interview guides were based on CFIR constructs of intervention characteristics, inner setting, and outer setting, as well as the Framework for Reporting Adaptations and Modifications-Enhanced (FRAME) for classifying intervention modifications.^22,24,25^ Interviews were audio-recorded and transcribed. Recruitment stopped when data saturation was achieved (i.e., additional interviews yielded no new insights).

### Data Analysis

For surveys, we calculated the mean (SD) for each 4-item measure. Higher scores indicate greater feasibility, acceptability, and appropriateness.^23^ We used independent samples t-tests to calculate mean differences between pre- and post-implementation respondents.

We analyzed qualitative data in MaxQDA software using thematic analysis. For deductive themes (e.g., CFIR constructs^22^, FRAME^24,25^), we described modifications to the intervention, including by whom modifications were made, what was modified, at what level of delivery, and nature of the content modification. Additionally, we identified inductive themes, or those arising from the data, using the following steps: 1) two researchers (among EY, LN, SR) read each transcript to become familiar with the text and to develop initial codes, 2) two researchers independently coded three transcripts iteratively and discussed disagreements to refine the codebook, and 3) the final codebook was determined by consensus. Inductive and deductive codes were applied to all transcripts using consensus coding (two coders independently coded each transcript and resolved discrepancies by consensus). Code summaries were developed that described the breadth and depth of each code, and final themes and subthemes were developed to create a cohesive message. These steps ensured all transcripts were coded by two researchers, maintaining rigor and reliability throughout the coding process.^26^

## Results

### Adult Oncology Program Pilot

#### Quantitative Results

We enrolled 33 healthcare providers to participate in the pre-(n=16) or post-implementation (n=17) questionnaires. Participant characteristics are summarized in Table 2.

**Table 2.** Participant characteristics (n=48)

| **Characteristics** | **Adult oncology setting**<br>**(n=33)**<br>**No. (% of total)** | **Pediatric oncology**<br>**setting (n=15)**<br>**No. (% of total)** |
| --- | --- | --- |
| Age, years (SD) | 40.7 (10.3) | 47.0 (9.0) |
| Gender |  |  |
| Female | 22 (66.7) | 11 (73.3) |
| Male | 11 (33.3) | 4 (26.7) |
| Race |  |  |
| White | 23 (69.7) | 11 (73.3) |
| Asian | 5 (15.2) | 4 (26.7) |
| Black/African American | 3 (9.1) | 0 (0.0) |
| Mixed race/ Some other race | 1 (3.0) | 0 (0.0) |
| Prefer not to answer | 1 (3.0) | 0 (0.0) |
| Hispanic | 3 (9.1) | 3 (20.0) |
| Provider Type |  |  |
| Physician | 21 (63.6) | 12 (80.0) |
| Advanced Practice Provider | 12 (36.4) | 1 (6.7) |
| Social Worker | 0 (0.0) | 2 (13.3) |

At pre-implementation, mean (95% confidence interval) appropriateness score for the BPA was 4.27 out of 5 (3.92, 4.61); mean (95% CI) acceptability score was 4.22 (3.83, 4.60); and mean (95% CI) feasibility score was 4.34 (4.03, 4.65). At 3 months post-implementation, mean (95% CI) appropriateness score for the BPA was 4.23 (3.86, 4.60); mean (95% CI) acceptability score was 3.92 (3.40, 4.44); and mean (95% CI) feasibility score was 4.33 (4.04, 4.62). There were no significant pre/post differences in mean scores for the BPA for each measure (p>0.05).

Audit data from the intervention demonstrated that 143 eligible patients were seen from April-October 2020 (Figure 3). Prior to implementation of the EHR-enabled intervention, the base rate of oncofertility screening was 30%, and after three months of pilot testing, the rate increased to 51% (n=72). Eight percent (n=11) were referred for oncofertility consultation; 100% of these completed a consultation (1 via telehealth) within a median of 4 days (range 2 to 14 days). Another 43% (n=61) had the following valid reasons for not pursuing fertility consultation: patient declined to address, already referred, poor prognosis, patient currently on treatment. Among the remaining 49% of patients, 11 (8%) were deferred to a future visit, and 60 (41%) had the BPA dismissed during the provider visit.

**Figure 3.**
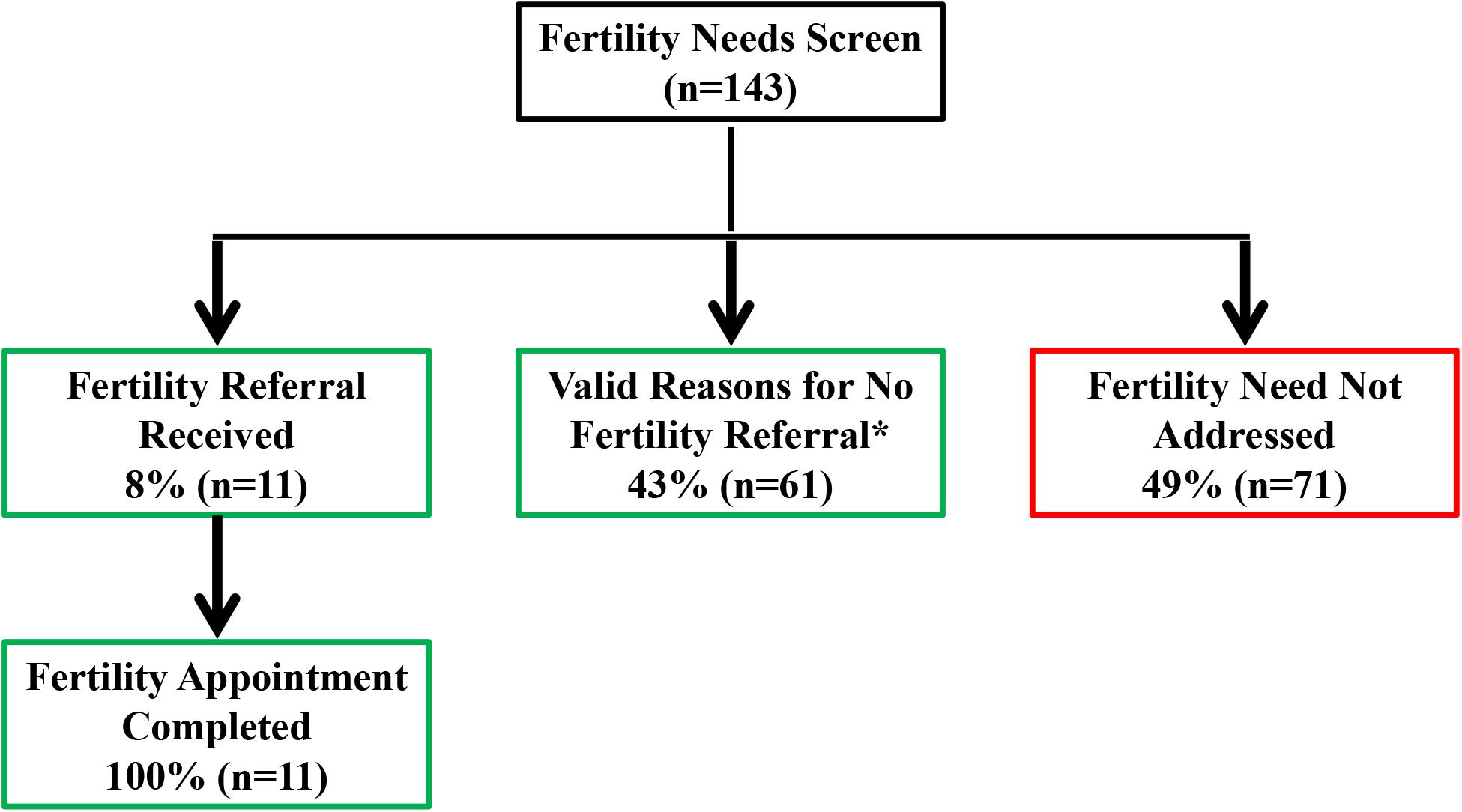
Electronic health record-enabled multi-component intervention for oncofertility care pilot data from an adult oncology program. Number of patients from Breast, Urology, Hematology Disease Teams April to October 2020 who receive goal-concordant care (green) or goal-discordant care (red). *Valid reasons for not referring include the following: Already referred, patient declines to address, patient currently on treatment, poor prognosis.

#### Qualitative Results

We conducted six interviews with providers from adult oncology and fertility teams. Overall, providers reflected positively on using the EHR-enabled screening and referral pathway in their current practices: *“I think that it’s well designed, and I’ve had a really good experience with it. So I’ve been very happy it exists*.*”* (Breast Team Physician). Advantages of the intervention included reminding providers to discuss fertility preservation and facilitating the referral process: *“That was always my concern is I never knew who to call, consult, or who to get Reproductive Medicine involved on. I think if it was kind of automated, that’s perfect*.*”* (Hematology Team Physician).

Table 3 highlights key themes regarding requested content and context modifications and potential solutions. For the content, providers wanted additional BPA reasons for not ordering a referral where currently they would dismiss the BPA because they could not find a valid reason. On the referral order, providers suggested not requiring completion of the anticipated treatment field because the plan of care may not yet be known or within the provider’s scope of care for ordering the referral, e.g., surgical oncologist. Context modifications included changes to the format and the personnel. For format, multiple providers recommended changing to the timing of initial BPA reminder to a patient’s second, rather than new oncology, visit, because the first visit is often too overwhelming to address fertility discussions. For the intervention personnel, some providers were interested in having the BPA fire for nurses, in addition to firing for physicians and APPs, as nursing may assist with reminding physicians in settings that lack APPs.

**Table 3.** Adult oncology program pilot qualitative interview feedback on content and context modifications to the EHR-enabled Best Practice Advisory screening and referral pathway

| Target | Content Modifications | Rationale – Illustrative Quotations | Potential Solution |
| --- | --- | --- | --- |
| BPA: Consult already ordered | Adding Elements | “It [the BPA] did pop up on one of my patients the other day, and I hadn't had that before where I looked in the chart and like the consult was already ordered. And I assume ... by the surgeon, and you know, I kind of wanted to make sure ... that I wasn't just kind of ignoring the fertility concerns and I wanted there to be a way that I could say, you know, consult is already happening or something like that.” (Breast Team Physician) | Add reason for not ordering referral: consult already ordered |
| BPA: Remind on next visit | Adding Elements | “If there was a way like instead of ordered, you know, order there was like, you know, a button for next time like or, you know, remind our next visit, because that would be helpful.” (Breast Team APP) | Add reason for not ordering referral: address next visit |
| Referral order: Anticipated Treatment Course field | Tailor/ Tweaking/ Refining | “The hard thing about the anticipated treatment courses sometimes we just don't know.” (Breast Team APP)<br>“I'm the surgical person, right. And so, I'm putting orders in, and I'm just guessing what the medical oncology treatment is, it's not really appropriate.” (Breast Team APP) | Remove as a requirement<br>Change to “potential treatment course” |
| Referral order: Medical provider contact information field | Removing Elements | “I would remove medical provider because yeah, you can just call whoever made the order.” (Breast Team APP)<br>“So I mean, because typically what you see if you see the referral, you know who it's from. So that just could be one thing you could potentially think about taking out.” (Hematology Team Physician) | Remove field |
| Target | Context Modifications | Rationale – Illustrative Quotations | Potential Solution |
| BPA: Fire during patient visit rather than during pre-charting | Format | BPA firing when providers were pre-charting and not in real time when they see patients: “I'm doing, most of, my notes are done at home...if you're not with the patient, and you're not discussing it with them at that time, right.” | Add criterion for patient to have arrived before the reminder fires |
|  |  | *We just said that'll be their next visit.”*<br>(Hematology Team Physician) |  |
| BPA:<br>Remind at<br>second visit | Format | • *“Especially when I'm very first seeing the woman, it's very overwhelming, right? And so, at first they'll might say, you know what, I don't want to even think about that – I'm just wanting to do this right now. And so, they're saying no. It's sometimes that second visit where they've had time to kind of think about things, and I think that's the, that's my problem with it is that it, it makes it only on their very first meeting. And especially on the surgical side where we're meeting them literally after they've been biopsied. And they found out it's so fresh, there's a lot of emotions... I'm not only doing more work up done in oncofertility, but I'm also talking about you know, Social Work, Psychology and so on. And again, it's a lot of things. And when somebody gets diagnosed with cancer, it's already scary enough. And they're walking out with, you know, 10 referrals and 20 tests to do. That's, that's a lot.”* (Breast Team APP) | • Add reason for not ordering referral: address next visit |
| BPA: Add nurses to providers eligible for the reminder | Personnel | • *“I mean, so most of us in hematology, we don't have APPs. So then, you know, we rely a lot on nurses to assist us. So, in that regard, then the nurse, maybe don't make it for an MA but make it maybe make it for the nurse and also the pop up so that would potentially, they could have a reminder to so they can always remind the clinicians.”* (Hematology Team Physician) | • Add additional personnel category, i.e., nurses, to BPA specification |

### Evaluation of Intervention Fit across Additional Oncology Settings

#### Qualitative Results

We interviewed 15 healthcare providers from Southern California academic and community oncology and fertility programs. These providers included physicians, APPs, and social workers (Table 2). Key themes regarding fit of the EHR-enabled multi-component intervention to various clinical contexts are presented below organized by CFIR domains.^22^

#### Intervention Characteristics

Key themes related to intervention characteristics focused on the relative advantage, adaptability, complexity, and cost of the intervention. The overwhelming majority of providers and social workers expressed that having the intervention would be a relative advantage to their current processes in terms of screening, referral, and patient access to telehealth fertility counseling: *“It’s like people aren’t going to fall through the cracks… this seems like an ideal state to work towards*.*”* (Social Worker). A fertility provider stated, *“I think it’s very doable to do the counseling over a video… if at the end of that fertility counseling visit, they feel satisfied… then I save them a trip coming in*.*”* Discussions about the adaptability of the screening/referral pathway centered around when and how many times and when the BPA should fire as well as the modifying the build for different instances of EPIC and additional EHR systems. Providers were supportive of the BPA screening and referral pathway build for their respective EHR systems (i.e., EPIC and Cerner), as both have similar functionalities.

Complexity involved referral orders potentially going to multiple fertility practices, including outside versus embedded programs: *“You’re expecting all your referrals go to one practice…when in actuality, … oncologists don’t always refer to me, they refer to other REIs as well*.*”* (Fertility Physician). Discussions about cost of implementing the intervention related to time and money needed to build the tool in the EHR system and the relative advantage of modifying an existing build: *“In the community hospital world, they’re about time and money… as simple as you can make it for them…*.*The key would be how do we make it a really quick seamless process*.*”* (Oncologist).

#### Outer Setting

Overall, providers felt that external policies and incentives prioritizing fertility care would help motivate implementation of the intervention. For example, one provider stated: *“I would think the US News rankings would* … *So I think that’s a motivator*.*”* (Fertility Physician). Another provider felt that making oncofertility care a target would be beneficial: “*If it was a hard measure of target for everyone’s performance… I think if it was made a target, I think there would be incentive*.*”* (Fertility Physician).

#### Inner Setting

Discussions of how to implement the intervention in their own context focused on the implementation climate and readiness for implementation. Most providers expressed that having a goal and quality metrics are important for oncofertility screening and referral: “*I think people like to have goals… and I think that if that’s established internally and agreed upon, then I think that they’ll rally behind*.*”* (Oncologist). Another viewpoint focused on the shared receptiveness of those who are delivering oncofertility care: *“I think it’s good, but it’s only as good as the person that’s clicking the box…And if, if the provider chooses to skip it they, they can cause like I said, not all providers are, are equally supportive*.*”* (Fertility Physician).

In terms of readiness for implementation, all sites expressed the need for leadership/administration engagement and buy-in to support changes to oncofertility care delivery: *“My concern would be more like getting the powers-that-be you to, like, agree to this system, because it would be such a large number of referrals across the board*.*”* (Social Worker). Some providers shared the value of getting the leadership involved prior to implementing the EHR-enabled intervention: *“Building something automated is-I have to go through the right channels. So that’s like the first hurdle there*.*”* (Oncologist).

## Discussion

Through the lens of implementation science frameworks (CFIR^22^ and FRAME^24,25^), we conducted a mixed-methods pilot study to assess the acceptability, appropriateness, and feasibility of an EHR-enabled multi-component intervention for oncofertility care in young cancer survivors at one adult oncology program as well as to assess needed adaptations to this intervention. Additionally, we explored key themes regarding intervention fit to various clinical contexts at seven additional oncology and fertility programs in Southern California. These adult and pediatric programs included both community and academic programs and represented both embedded and outside fertility programs. For the adult oncology program pilot, the intervention was rated as highly appropriate, acceptable, and feasible, and provider feedback identified modest context and content modifications to the EHR-enabled screening and referral pathway. Overall, the intervention resulted in appropriate referrals and oncofertility consults and improved data capture of oncofertility care.

While the EHR-enabled multi-component intervention increased the rate of goal-concordant oncofertility care delivery and was positively rated by providers both pre- and post-implementation at the adult oncology program, approximately half of eligible patients did not have them adequately addressed. To address the moderate number of reminders that were dismissed, the BPA can be modified to include appropriate reasons such as a snooze to the next visit and not allow the option for dismissal. Tailoring is pragmatic at the institutional level rather than the clinic level to avoid building redundant BPAs within one EHR system. This renders user-centered design, sampling enough end users, post-implementation feedback, and consideration of the oncofertility care delivery literature to inform which modifications to incorporate. For example, a few physicians discussed adding nurses to the reminder pool, consistent with ASCO practice guidelines recommending involvement of nurses in oncofertility education.^5^ Yet many reports from the nursing perspective demonstrate that oncofertility education is outside their scope of practice due to lack of knowledge and comfort with this topic,^7,27^ rendering our not modifying the intervention. Taken together, more work is needed to appropriately identify the targets and timing of the BPA tool, so that providers feel confident to engage in oncofertility discussions with their patients, instead of dismissing the BPA.

Disseminating the EHR-enabled multi-component intervention requires identifying adaptations and assessing readiness of new clinical settings for the intervention.^28,29^ The use of CFIR^22^ and FRAME^24,25^ allowed us to systematically assess and identify facilitators, barriers, and adaptations to scale up the EHR-enabled intervention for diverse community, academic, pediatric, adult oncology and fertility settings. Across these diverse settings, providers agreed the intervention would be advantageous for oncofertility care delivery and were able to identify key themes associated with intervention fit within their existing clinical contexts. Existing HER systems can easily facilitate these intervention modifications and allow for tailoring by clinical site.

One limitation of our study is the inclusion of oncology programs with existing EHR systems and fertility clinic referrals sites; hence, our findings may not be generalizable to other settings, such as rural oncology clinics without EHR platforms or existing fertility referral pathways. However, our EHR-enabled intervention has the potential to be adapted to settings without EHR systems through a paper-based screening and referral system. Additionally, our sampling strategy for healthcare providers was purposive and not random and may result in selection bias if the healthcare providers who participated in our study have an interest in oncofertility care.

## Conclusions

In summary, our findings highlight that the EHR-enabled multi-component intervention for oncofertility care delivery was highly acceptable, appropriate, and feasible as well as effective at improving screening for oncofertility needs among young cancer patients. Further, we identified potential barriers, facilitators, and adaptations to the EHR-enabled intervention that may be necessary to improve oncofertility care of patients and scale up the intervention across diverse adult and pediatric oncology settings.

## Data Availability

The data that support the findings of this study are available from the corresponding author, [SADR], upon reasonable request.

## Acknowledgements

We wish to thank Kristopher Brodsho for guidance on EHR capabilities. We would also like to thank healthcare providers and clinical staff at all sites for their contributions to this study.

## Author contributions

Design and conceptualization (HIS, PA, TH, SA, LT, SADR); Data acquisition (EHY, AD, LN, TH, HIS, SADR); Data analysis (EHY, AD, LN, BNK, HIS, SADR); Data interpretation (all authors); Manuscript preparation and revision (all authors); Manuscript final approval (all authors)

